# Intravenous iron and iron deficiency anemia in gastrointestinal cancer patients: a systematic review

**DOI:** 10.1101/2023.09.20.23295822

**Authors:** Shankavi Nandakumar, Navreet Singh, Alliya Remtulla Tharani, Christine Brezden-Masley

**Author notes:** Corresponding author: Christine Brezden-Masley, Department of Medicine, Mount Sinai Hospital, Toronto, ON, Canada. **Email addresses:**.

## Abstract

**Background:** Iron deficiency anemia (IDA) is a prevalent hematological complication associated with gastrointestinal (GI) cancers due to an increased loss of iron and decreased iron absorption. We estimated the efficacy of parenteral iron on hemoglobin levels, blood transfusion needs and overall quality of life in patients with GI malignancies.

**Methods:** In this systematic review, we used PubMed, Cochrane, EMBASE, CINHAL and Scopus to conduct an electronic search from January 1, 2010 to March 24, 2022 with no language or study design restrictions. Studies were included if they discussed IDA, GI neoplasms, use of iron supplementation (with or without erythropoietin-stimulating agents [ESAs]), defined anemia and had a patient population of adults. Studies were excluded if were published before 2010. We assessed the efficacy of parenteral iron in comparison to other iron supplementation methods when treating IDA in GI cancer patients. The Cochrane Risk of Bias Tool 2 (RoB 2) and the Risk Of Bias In Non-randomized Studies of Interventions (ROBINS-I) assessment tools were used to assess the quality of the included studies. Moreover, the Cochrane Effective Practice and Organization data collection form was used to collect pertinent study information.

**Results:** Our search yielded 3,156 studies across all databases. With the exclusion of duplicates, ineligible study designs, as well as studies that did not pass abstract and full-text screening, 17 studies were included in our final analysis (4 randomized control trials; 13 non-randomized studies). Of the 13 studies evaluating hemoglobin (Hgb) response, seven studies found an increase in Hgb levels when patients were treated with IV iron. The 8 studies evaluating red blood cell (RBC) transfusion rates found no significant differences in RBC transfusion needs when treated with IV iron. Studies analyzing health related outcomes typically found an increase quality of life and decreased post-operative complications.

**Discussion:** This review reveals the improved outcomes of IDA in GI cancer patients treated with IV iron instead of other iron supplementation methods. Timely diagnosis and appropriate IDA management can greatly improve quality of life in this patient population, especially if myelosuppressive chemotherapy is required.

Our systematic review presents some limitations due to heterogenous interventions in the randomized control trials, the varying time points of data collection in each study, and the use of small sample sizes.

## Introduction

Luminal gastrointestinal (GI) cancers (i.e., cancers of the esophagus, stomach, large and small intestine) represent 26% of global cancer incidence, and 35% of all cancer-related deaths [1]. A common hematological complication associated with luminal GI cancers both at diagnosis and during treatment is anemia, defined by the World Health Organization (WHO) as hemoglobin (Hgb) of less than 12 g/dL in women, and less than 13 g/dL in men [2,3]. The cause of anemia in cancer patients is multifactorial in nature, and can be attributed to comorbidities such as bleeding, hemolysis and nutritional deficiencies [4], such as iron deficiency. The prevalence of iron deficiency anemia (IDA; defined as anemia associated with iron deficiency) in GI cancer patients ranges from 7% to 42% [5]. The etiology of IDA in patients with GI cancer can be attributed to increased loss of iron (i.e. bleeding) and decreased iron absorption [6].

In GI cancer patients, IDA is associated with poor prognosis, fatigue, and poor quality of life (QoL) [7]. Targeted treatment of anemia in patients can both improve prognosis and QoL [8]. Common treatment approaches in this patient population include red blood cell (RBC) transfusions, erythropoietin stimulating agents (ESA), and iron supplementation. However, despite available treatment options, approximately 60% of patients with anemia do not receive any treatment [2]. Potential reasons for patient undertreatment may include: poor optimization of care across the patient journey, unclear guidelines, and inadequate testing for anemia and IDA [2].

Treatment of anemia with RBC transfusions should be used cautiously as use is associated with increased risk of morbidity and mortality in cancer patients[9]. ESAs offer a means of reducing the need for RBC transfusions but, only 30-75% of patients may respond to treatment and use may increase the risk of thromboembolic events [9]. Furthermore, intravenous/parenteral (IV) iron alone or in combination with ESAs presents itself as an effective treatment for anemia, while also reducing the need for RBC transfusions [10]. Oral iron supplementation is also commonly prescribed to address IDA in the cancer population; however, evidence suggests oral iron does not reduce the risk of RBC transfusion, most likely due to either malabsorption of oral iron, non-adherence and also slow bioavailability and repletion [11].

The 2018 European Society for Medical Oncology (ESMO) clinical practice guidelines recommend that RBC transfusions only be used in patients with severe anemia-related symptoms, and ESAs only be employed when patients undergoing chemotherapy have had their iron deficiency corrected. Additionally, the American Society of Clinical Oncology (ASCO)/American Society of Hematology (ASH) recommends that ESAs be offered only to patients whose cancer treatment is not curative in intent and with Hgb <10 g/dl [12]. However, this recommendation is based on a lack of evidence indicating whether a particular patient population receiving ESAs is at greater or lesser risk of harm particularly in terms of progression/reoccurrence and overall survival [12]. Currently, there are no specific guidelines that can aid physicians in making decisions about treating IDA in patients suffering from GI cancers and, existing guidelines do not provide recommendations regarding the use of parenteral iron supplementation with/without ESAs, and do not discuss when patients are to receive treatment. In addition, existing systematic reviews predominantly focus on the use of parenteral iron supplementation for treating chemotherapy-induced anemia, and the addition of parenteral iron to ESAs in cancer patients more broadly [11,13,14]. To our best knowledge, no systematic review exists to date, that examines the use of IV iron in patients with GI cancers with respect to when patients are being diagnosed with IDA, when they are being treated, how they are optimally treated, and the benefits of treatment.

Therefore, given the heightened prevalence of IDA in this patient population, the purpose of this systematic review is to evaluate the use of IV iron to treat IDA in patients with GI cancer.

## Methods

This systematic review was performed following the Cochrane Training Handbook guidelines, as well as the Preferred Reporting for Systematic Reviews and Meta-Analyses (PRISMA) 2020 checklist [15,16]. This review was not registered, and the review protocol is not available.

### Search Strategy

A search using the following databases: PubMed, Cochrane, EMBASE, CINHAL, and Scopus was conducted. The search terms included but were not limited to: “iron deficiency”, “anemia”, “gastric cancer”, “ESA therapy*”, “intravenous iron” and “iron studies”. Randomized control trials (RCTs), systematic reviews, observational studies, case studies, and cohort studies, from January 2010 to April 2022, with no specified language restrictions were retrieved.

### Study Selection

Studies were reviewed if they included the following: 1) iron deficient anemia, 2) gastrointestinal neoplasms, 3) iron supplementation alone (i.e. intravenous iron or parenteral iron, oral iron) or in conjunction with ESAs, 4) defined anemia and the symptoms associated with anemia, and 5) an adult (≥ 18 years of age) population. Additionally, literature published before 2010 was excluded from this systematic review as the use of IV iron became readily available for the correction of anemia in 2010 [17].

The web-based software, Covidence^TM^, was used by two authors (SN and NS) to screen studies and extract data[18]. A standardized eligibility checklist was used to screen the title, abstract and full-text of studies; removing ineligible studies throughout the process. Any conflicts that arose were discussed and resolved between the authors (SN and NS).

### Data Collection and Quality Assessment

The Cochrane Effective Practice and Organization data collection form was used to curate a standardized extraction sheet to collect the study information needed from each extracted article [15].

The Cochrane Risk of Bias Tool 2 (RoB 2) was used to assess risk of bias among eligible RCTs [19]. RCTs were assessed and given an overall judgement of low risk-, some concerns-, or high risk of bias [19]. Moreover, the Risk Of Bias In Non-randomized Studies of Interventions (ROBINS-I) tool was used to assess the quality of non-RCT studies [20]. The non-RCT studies were given an overall judgement of low, moderate, serious, or critical risk of bias [20]. The risk assessment for this study was completed individually by reviewers (SN and NS). Visualization of risk-of-bias assessments were generated using the *robvis* online tool [21].

### Statistical Analysis

Statistics used in the study were expressed as means, medians, standard deviations (SDs), interquartile ranges (IQRs), and 95% confidence intervals (CI) for any relevant study variables.

## Results

Seventeen studies published between January 1, 2010 and March 24, 2022 met the inclusion criteria and were included in the final analysis. A summary of the screening process can be found in Figure 1. Of these studies, four RCTs compared IV iron supplementation to standard of care or compared two IV iron interventions to one another. The remaining 13 studies were of non-randomized design (8 retrospective, 5 prospective).

**Figure 1.**
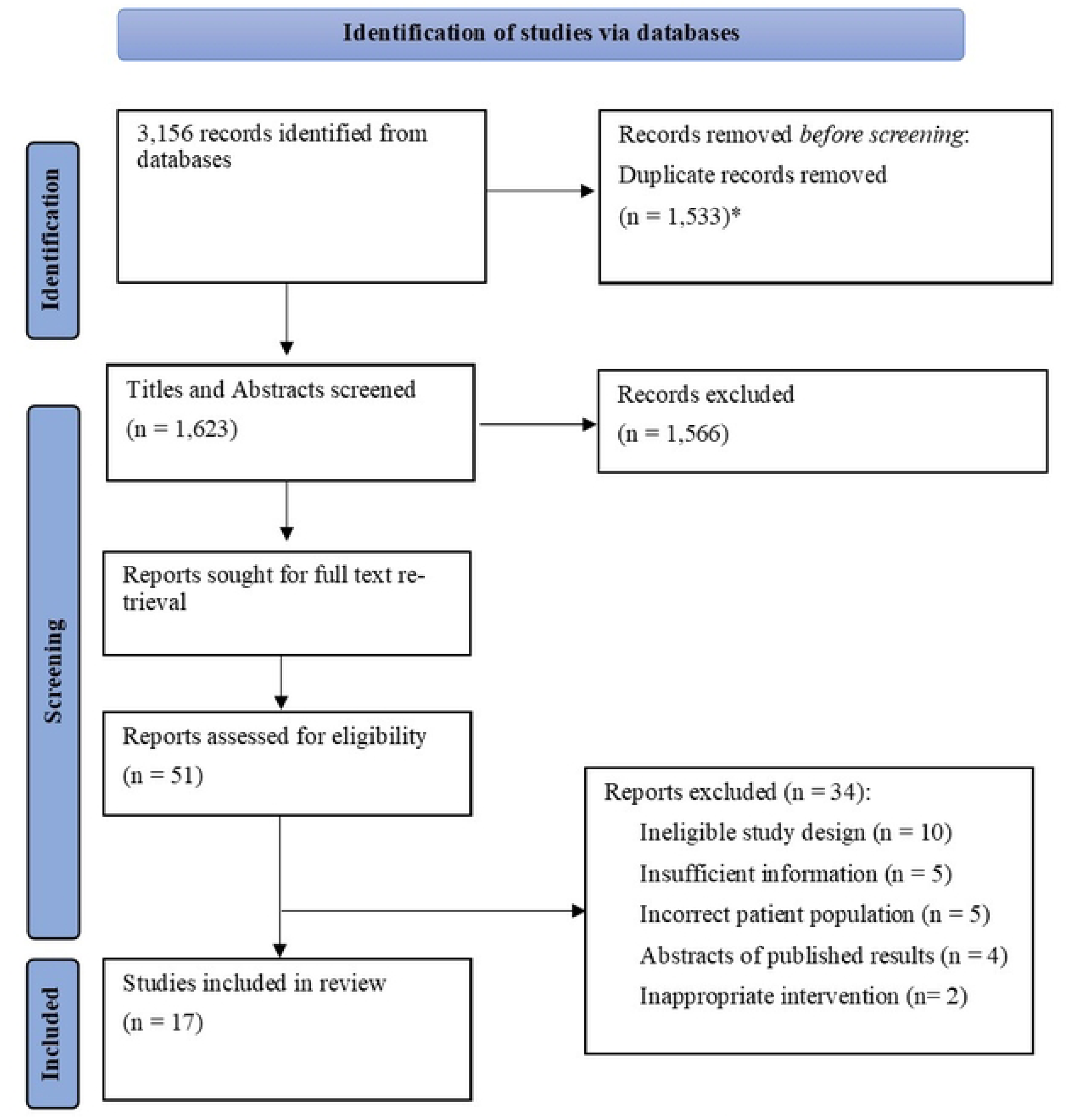
PRISMA Diagram for study identification, screening, and inclusion. * Duplicates removed by software program.

All studies included a comparator arm, except for Bojesen et al. [22], Lima et al.[23] and, Verhaeghe et al. [24] where all patients received IV iron. Of the RCTs, Keeler et al. [25] and Keeler et al. [26] compared ferric carboxymaltose (FCM) to oral iron, while Laso-Morales et al. [27] compared FCM to iron sucrose (IS), and Ng et al. [28] compared iron isomaltoside to standard of care. The comparator arms in eight studies [29–36] were patients who received no specific treatment or oral iron. Of note, Quinn et al.[37] compared the efficacy of a new anemia management intervention in which patients were given FCM, and then retrospectively compared to pre-intervention patients. IV iron treatment was provided preoperatively in 13 studies and postoperatively in four studies. A complete summary of the studies can be found in Table 1.

**Table 1.**
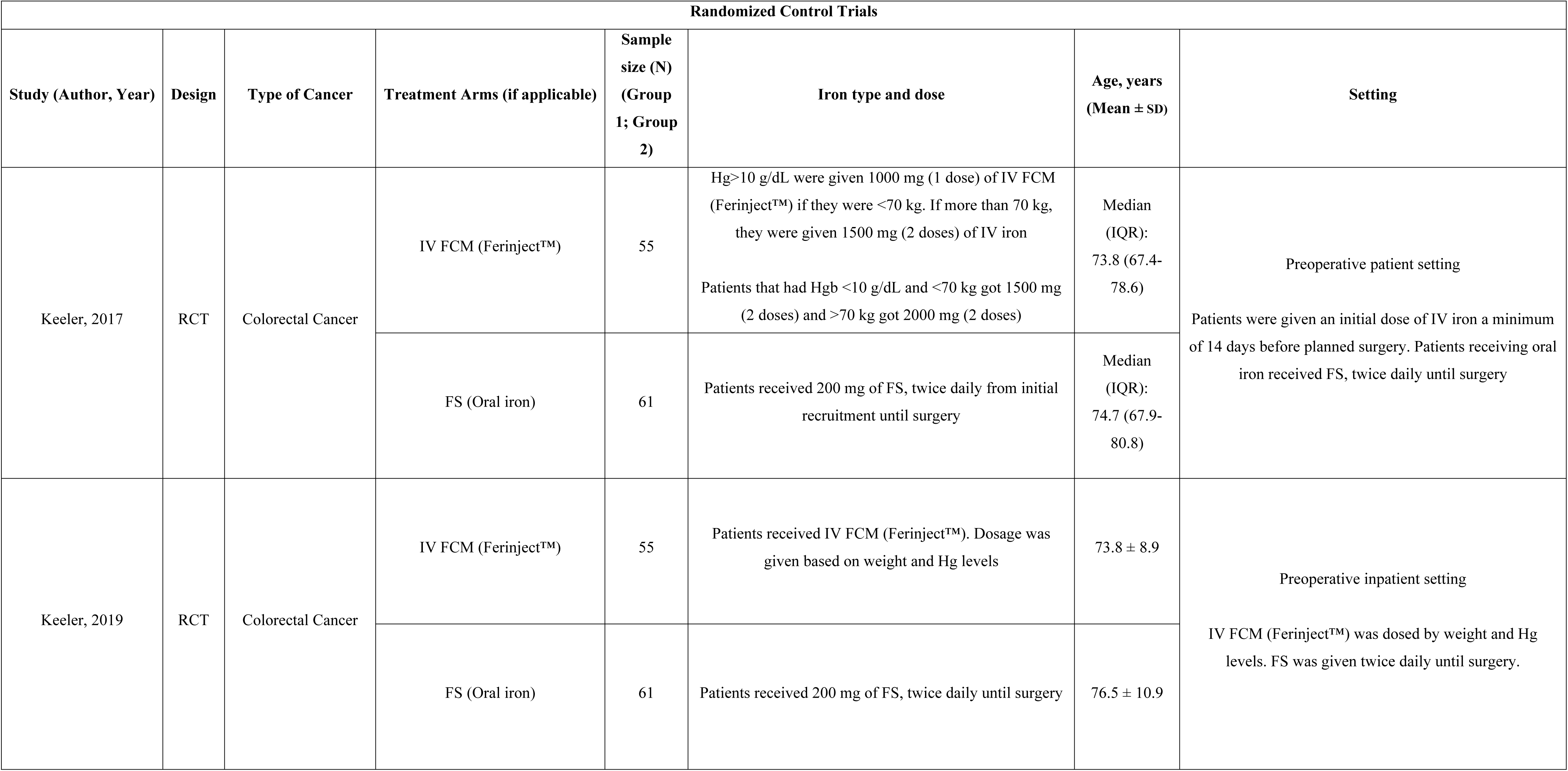
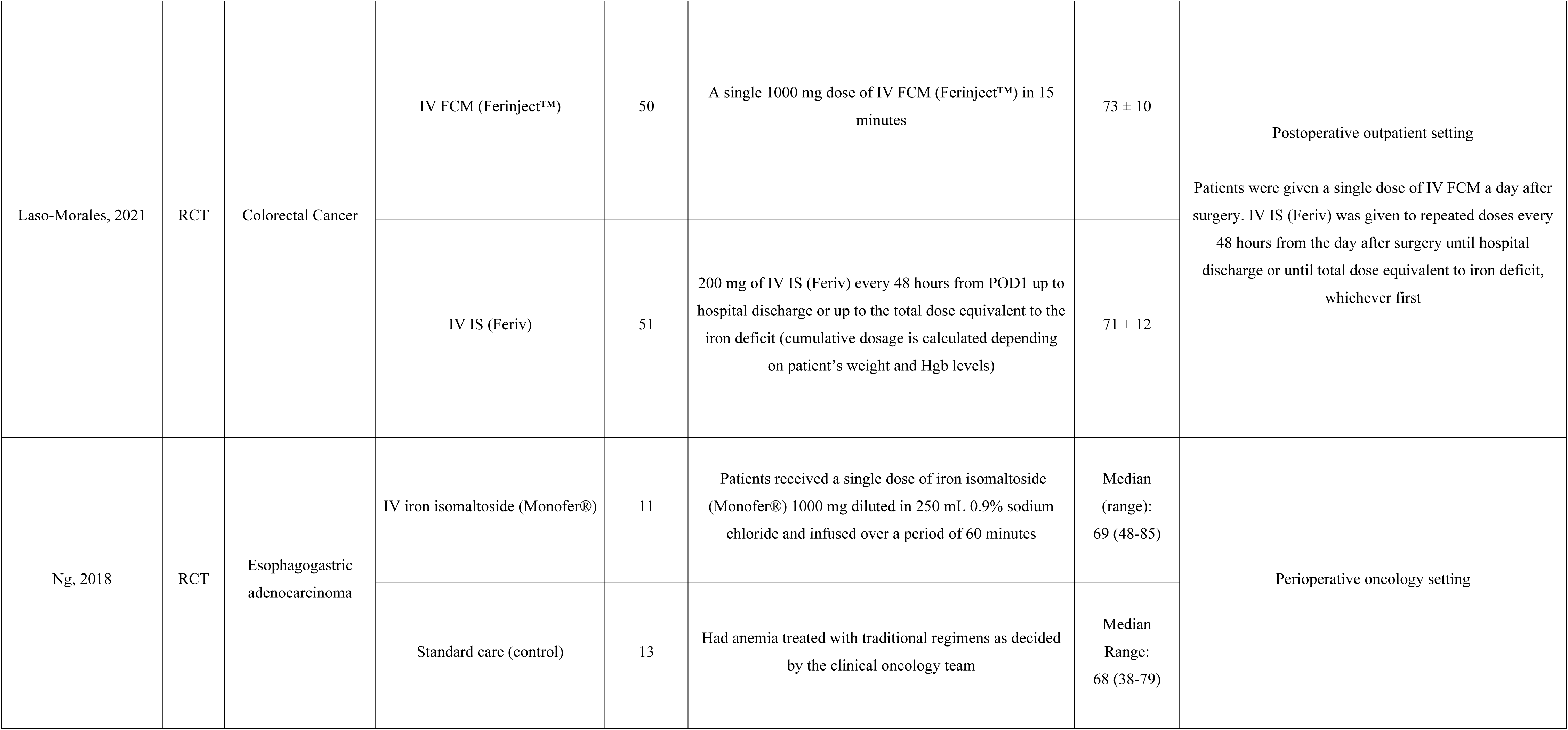
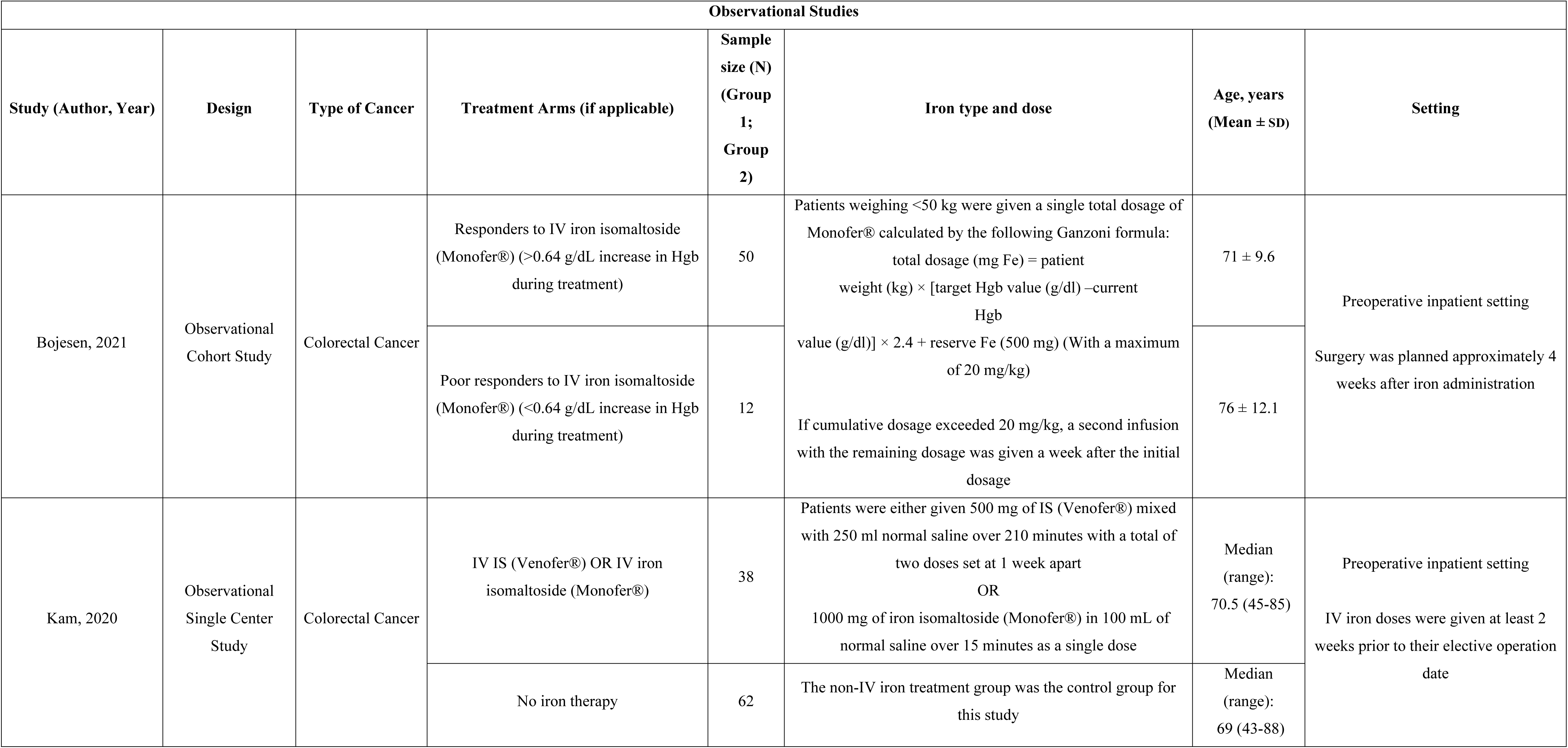
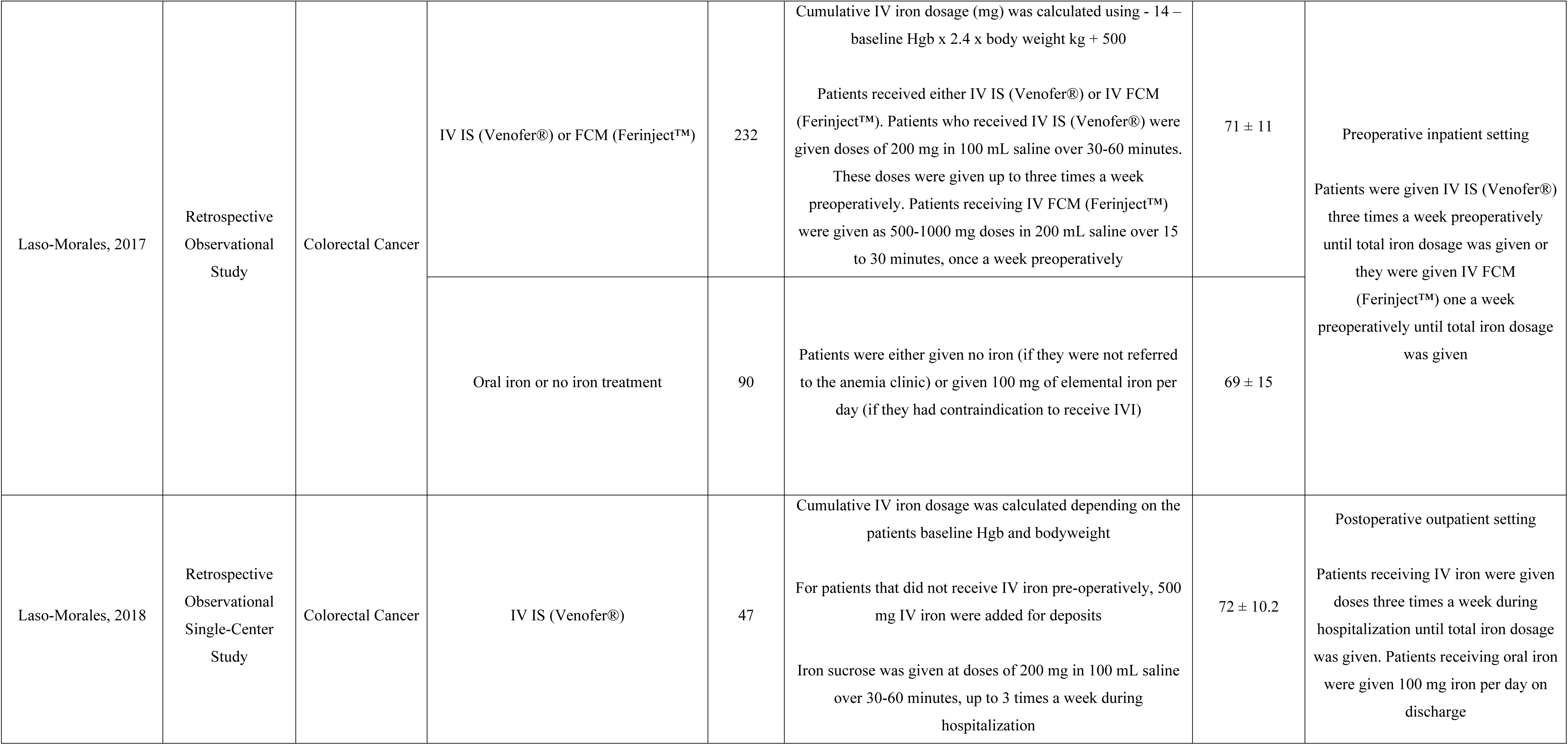
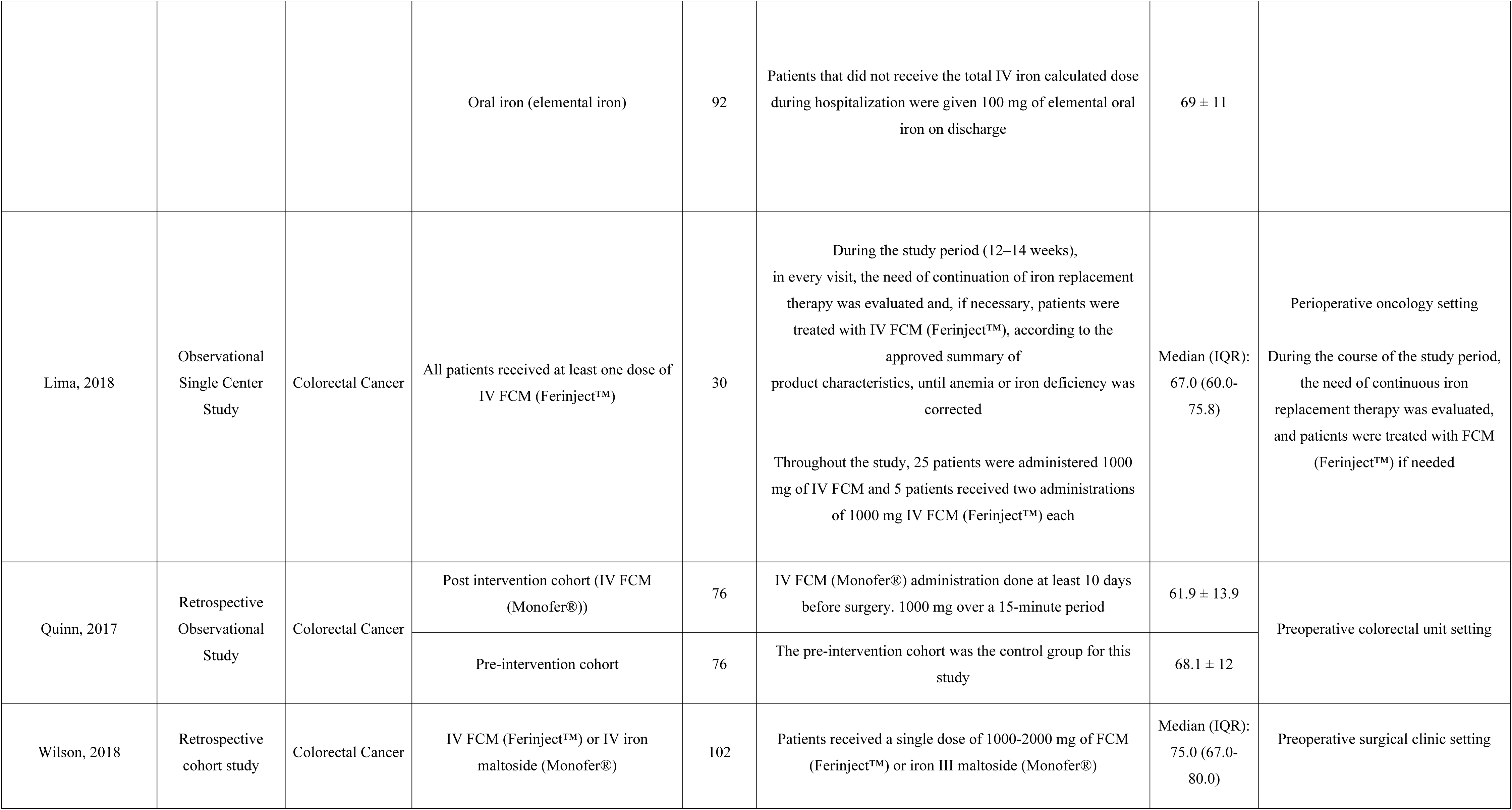
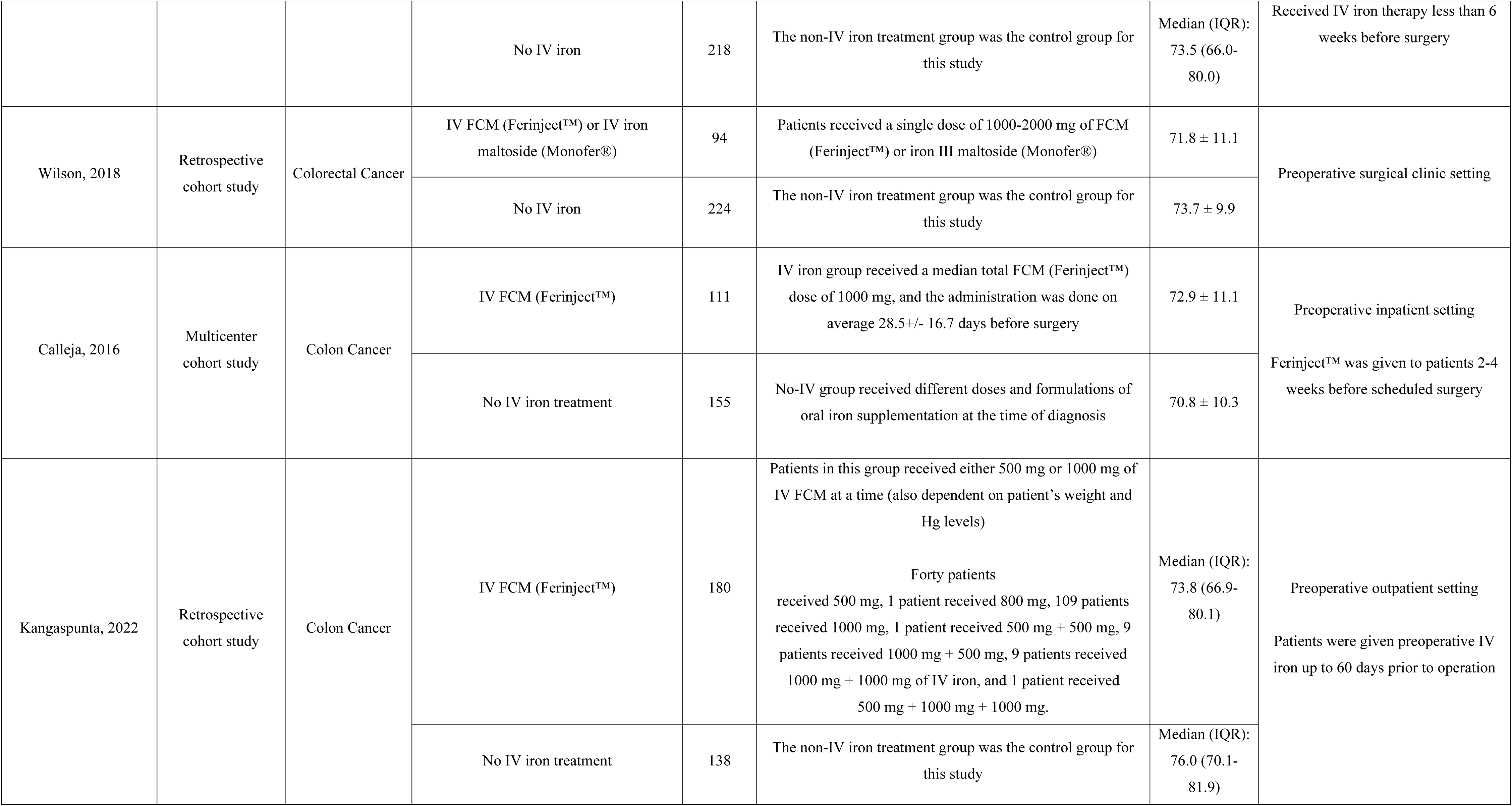
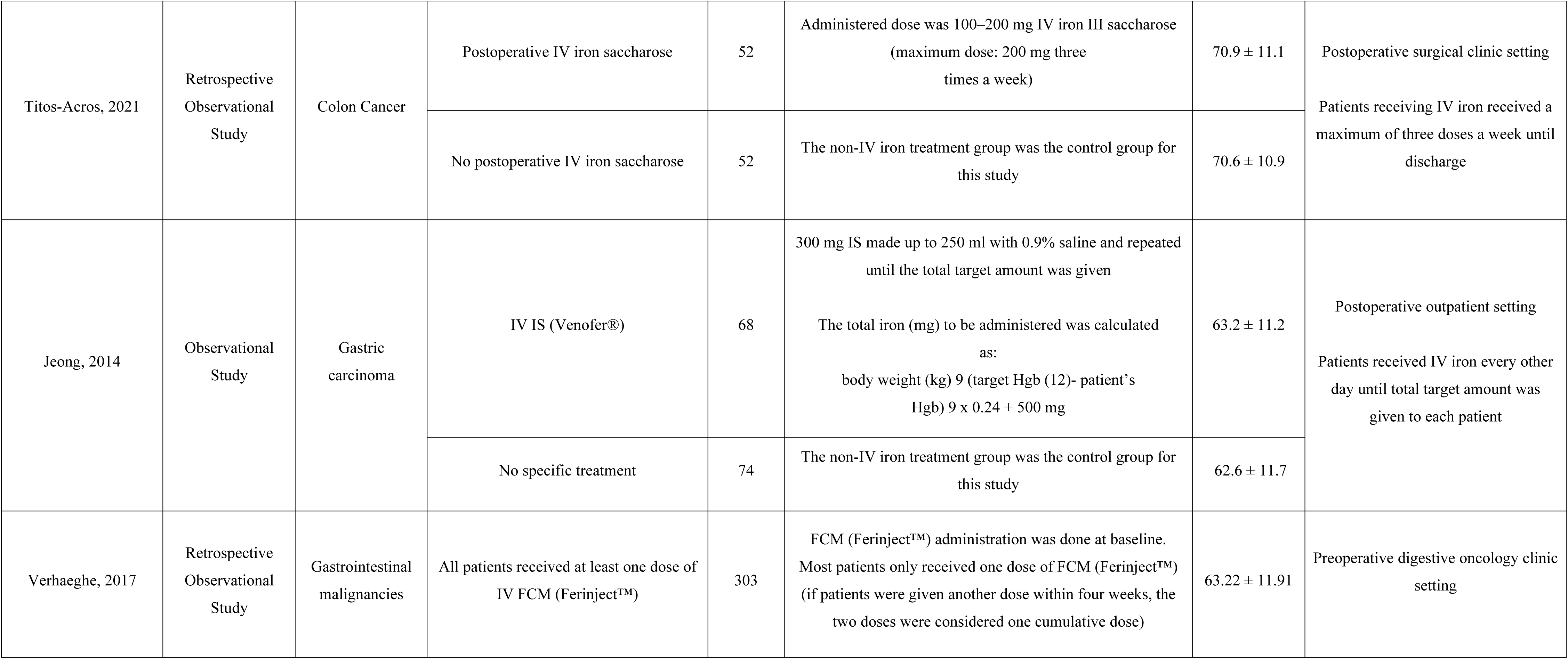
Demographic Characteristics of Included Studies.

**Table 2.** Main Results of Included Studies.

| Randomized Control Trials |  |  |  |  |  |
| --- | --- | --- | --- | --- | --- |
| Study (Author, Year) | Treatment Arms (if applicable) | Diagnostic Criteria | Outcomes of Interest | Results | Conclusion |
| Keeler, 2017 | IV FCM (Ferinject™)<br>vs.<br>FS (Oral Iron) | Hgb <11g/ dL in women<br>Hgb <12g/dL in men | Hgb levels | Increases in Hgb were higher in IV iron recipients (median 1.55 g/dL [IQR 0.93-2.58] vs. 0.50 g/dL [IQR -0.13 to 1.33]; p<0.001). Hgb levels were higher at surgery after treatment with IV iron than oral iron | IV iron did not reduce blood transfusion needs but was more effective than oral iron at treating preoperative anemia and iron deficiency |
|  |  |  | Blood Transfusion Rates | No difference in blood transfusion needs from recruitment to trial completion in terms of volume of blood administered (p=0.841) or number of patients transfused (p=0.470). |  |
|  |  |  | Iron parameters (Serum ferritin, TSAT) | Ferritin levels were significantly higher in the intravenous group at surgery (median 558 (IQR 330–1085) µg/l versus 27·5 (17–51·5) µg/l in oral group; P <0·001)<br>This same relationship was evident with TSAT levels at surgery (median 19 (16–29) and 9(5–14) % respectively; P <0·001) |  |
|  |  |  | Postoperative complications | There was no difference in grade of complication severity between groups from recruitment to outpatients (P =0·995) or in complication rate over the same period (P =0·305). The same was true for grade (P =0·083) and rate (P =0·091) of infective complications. |  |
|  |  |  | 90-day mortality | Ninety-day mortality rates were similar (2 and 3 deaths respectively; P =0·906). |  |
|  |  |  | Postoperative length of hospital stay | Postoperative length of hospital stay was 6 days for both groups (IQR 4–9 and 5–10 days for oral and intravenous groups respectively; P =0.950) |  |
| Keeler, 2019 | IV FCM (Ferinject™)<br>vs.<br>FS (Oral Iron) | Hgb <11g/ dL in women<br>Hgb <12g/dL in men | Hgb levels | Hgb levels were higher at surgery and in outpatients with IV iron however, no difference in blood transfusion use was seen | IV iron was more efficacious at improving QoL scores than oral iron |
|  |  |  | QoL | Eleven QoL components increased by a clinically significant margin in the IV iron group between recruitment and surgery with one component for oral iron. Fact-An scores were higher for the IV iron group. |  |
| Laso-Morales, 2021 | IV FCM (Ferinject™)<br>vs.<br>IV IS (Feriv) | Hgb <11 g/dL for all patients | Hgb levels | No between-group differences in mean change in Hgb from postop day 1 to postop day 30 (FC: 2.5 g/dL, 95% CI: 2.1-2.9; IS:2.4g/dL, 95% CI:2.0-2.8; p=-0.52) | Hgb increase was similar for both IV FCM and IV IS, but infection rate was higher in IS |
|  |  |  | Blood Transfusion Rates | No between-group differences in blood transfusion rates |  |
|  |  |  | Iron parameters (Serum ferritin, TSAT) | Both ferritin and TSAT levels were higher on the day of surgery than at the time of CRC diagnosis for FCM group |  |
|  |  |  | Postoperative complications | Infection rate was lower in the FCM (Ferinject™) group compared to IS (Feriv) group (9.8% vs. 37.2%). |  |
|  |  |  | Postoperative length of hospital stay | No between-group differences in length of hospital stay |  |

| Ng, 2018 | IV iron isomaltoside (Monofer®)<br>vs.<br>Standard care (control) | Hgb <12g/dL in women<br>Hgb <13 g/dL in men | Hgb levels | No significant change in Hgb between both groups with an increase in ferritin levels in IV iron group after cycle one of chemotherapy (Standard care: 116 ng/ml; IV iron 770 ng/ml, P<0.05). | IV iron improves QoL and ferritin levels |
| --- | --- | --- | --- | --- | --- |
|  |  |  | Blood Transfusion Rates | No difference in blood transfusion rates |  |
|  |  |  | Iron parameters (Serum ferritin, TSAT) | Ferritin showed a significant increase after chemotherapy cycle one in the group treated with IV iron 105 ng/ml to 1015 ng/ml (P<0.05) |  |
|  |  |  |  | Transferrin saturations increased above 20% in the intravenous iron group rising from 11.1% to 26.1% after cycle 1 ( P=0.196). Transferrin saturations never exceeded 20% in the standard care group but did rise from 11.9 to 19% after cycle three of chemotherapy. |  |
|  |  |  | QoL | IV iron group saw improvements in QoL |  |
|  |  |  | Anemia-specific QoL | IV iron group saw improvements in anemia-specific QoL. |  |
| Observational Studies |  |  |  |  |  |
| Study (Author, Year) | Treatment Arms (if applicable) | Diagnostic Criteria | Outcomes of Interest | Results | Conclusion |
| Bojesen, 2021 | Responders to IV iron isomaltoside (Monofer®) (>0.64 g/dL increase in Hgb during treatment) | Hgb ≤11.28 g/dL for all patients | Hgb levels | Hgb levels improved in patients over the duration of treatment: | Patients with severe anemia (Hgb<9.02 g/dL) had the largest increase in Hgb after 4 weeks. |
|  | vs.<br>Poor responders to IV iron isomaltoside (Monofer®) (<0.64 g/dL increase in Hgb during treatment) |  |  | Week 1: 0.77 g/dL (95% CI 0.52-1.03 g/dL, P<0.0001)<br>Week 2: 1.5 g/dL (95% CI 1.21-1.80 g/dL, P<0.0001)<br>Week 4: 2.13 g/dL (95% CI 1.71-2.55 g/dL, P<0.0001) | Patients with mild anemia (>10.31 g/dL) did not have an increase in Hgb during the treatment course. |
|  |  |  | Iron parameters (Serum ferritin, TSAT) | Poor responders had significantly higher ferritin levels at weeks 1, 2 and 4 (P = 0.02, P = 0.003 and P = 0.013, respectively) compared to responders.<br>Poor responders had a significantly lower increase in TSAT 1 week after initial treatment (−11%, 95% CI −18% to −4%, P = 0.0026) compared to responders.<br>No difference in serum iron was found between poor responders and responders at week 1, 2 or 4 |  |
|  |  |  | Postoperative complications | Twenty-one patients (34%) developed a postoperative complication within the first 30 days after surgery. Not responding sufficiently to treatment was associated with higher baseline Hgb (P = 0.049) and lower body mass index (P = 0.036) but did not remain significant after correction for multiple testing. |  |
|  |  |  | Postoperative length of hospital stay | The median length of stay was 3 days (IQR 3–6 days, range 1–36 days). |  |
| Kam, 2020 | IV IS (Venofer®) OR IV iron isomaltoside (Monofer®)<br>vs.<br>No iron therapy | Hgb <10 g/dL<br>Hgb <12 g/dL after recent transfusion | Hgb levels | No statistically significant difference in mean preoperative Hgb level between two groups. IV iron group had a higher median Hgb rise of 1.9 g/dL vs. 0.6 g/dL in non-IV iron (P<0.001). | Patients with severe anemia (Hgb<9.02 g/dL) had the largest increase in Hgb after 4 weeks.<br>IV iron can significantly increase Hgb levels in IDA patients before colorectal surgery and reduce RBC transfusions. |
|  |  |  | Blood Transfusion Rates | 8 patients in the IV iron group required blood transfusion compared to 30 in the non-IV iron group (P=0.006).<br>No significant difference in number of patients with red blood cell transfusion prior to IV iron (42.1% in IV iron and 41.9% in non-IV iron, p=0.987) |  |
| Laso-Morales, 2017 | IV IS (Venofer®) OR IV FCM (Ferinject™)<br>vs.<br>Oral iron or no iron | Hgb<13 g/dL for all patients | Blood Transfusion Rates | There was no difference in RBCT rates between anemic patients on IV iron and those on standard care (16% vs. 17%; p=0.865). | IV iron was more effective in treating preoperative anemia and reducing infection rate than standard of care |
|  |  |  | Postoperative complications | Infection rates were lower among IV iron treated patients (18% vs. 29%; p<0.05). IV iron did not reduce postoperative blood transfusions but was more effective than standard care in treating preoperative anemia. |  |
|  |  |  | Postoperative length of hospital stay | No significant differences in length of stay |  |
| Laso-Morales, 2018 | IV IS (Ferinject™)<br>vs.<br>Oral iron (elemental iron) | Hgb<12 g/dL for all patients | Hgb levels | Anemia was more prevalent among patients in the IV iron group (p=-0.027), despite greater increment in Hgb (2.0 ± 1.5 g/dL vs. 1.1 ± 1.2 g/dL; p=0.001). | Compared to standard of care, post-operative IV iron administration improved recovery of Hgb levels at 30 |
|  |  |  | Blood Transfusion Rates | 7 IV iron patients and 4 non-IV iron patients required postoperative transfusions. | days post-operative, without an increase in post-operative complications. |
|  |  |  | Postoperative complications | No differences in post-operative complications between the groups, and no IV iron-related adverse events. |  |
| Lima, 2018 | All patients received at least one dose of IV FCM (Ferinject™) | 1) Hgb ≤11 g/dL or a reduction ≥2 g/dL since the start of the current chemotherapy regimen<br>AND,<br>2) Ferritin <30 ng/mL and transferrin saturation (TSAT) <20%<br>OR,<br>Ferritin 30-800 ng/mL, and TSAT<50% | Hgb levels | Statistically significant increase in mean Hgb (10.3 vs 11.2 g/dL) | FCM (Ferinject™) was well tolerated and had a positive impact on treatment of IDA |
| | | | Iron parameters (Serum ferritin, TSAT) | Ferritin had a mean change of 646.7±455.7 ng/mL (p<0.001)<br>TSAT had a mean change of 6.7%±6.9% (p<0.001)<br>Serum iron had a mean change of 17.4±26.6 mg/mL (p<0.001) | |
| Quinn, 2017 | Post intervention cohort (IV FCM - Ferinject™)<br>vs.<br>Pre-intervention cohort | Hgb <11g/dL for all patients | Hgb levels | Mean day 3 post-operative Hgb levels were significantly lower in patients with uncorrected anemia (9.5 g/dL, p=0.004). | IDA management pathway resulted in improved perioperative Hgb levels with reduction in perioperative transfusion. |
|  |  |  | Blood Transfusion Rates | Postoperative transfusion rates were 38% in patients with uncorrected anemia, compared to 0% in corrected anemia and 3.5% in non-anemic patients. |  |
|  |  |  | Postoperative complications | There were seven postoperative transfusions in the entire cohort: two in non-anemic patients (3.5% transfusion rate), two in patients with iron deficiency (13% transfusion rate) and three in patients with uncorrected anemia from other causes (a 50% transfusion rate). |  |
|  |  |  | Postoperative length of hospital stay | The difference in length of stay between patients with corrected anemia and non-anemic patients was non-significant (p = 0.472). |  |
|  |  |  | Iron deficiency rates | Treatment resulted in a preoperative IDA correction rate of 60% (nine of fifteen). |  |
| Wilson, 2018 | IV FCM (Ferinject™) or IV iron maltoside (Monofer®) vs. No IV iron | Hgb < 12.0 g/dL in women<br>Hgb < 12.9 g/dL in men | 5-year OS | No significant differences in 1-, 3-, and 5-year OS between the IV iron and no IV iron groups | IV iron does not have a profound effect on long-term OS and DFS. |
|  |  |  | DFS | No significant differences in DFS between the IV iron and no IV iron group |  |
| Wilson, 2018 | IV FCM (Ferinject™) or IV iron maltoside (Monofer®) vs. No IV iron | Hgb < 12.0 g/dL in women<br>Hgb < 12.9 g/dL in men | Hgb levels | In the IV iron group, preoperative Hgb level was significantly increased compared to the non-IV iron group (0.65 mmol/L vs. 0.10 mmol/L, p<0.001). | IV iron leads to optimization of preoperative Hgb levels |
|  |  |  | Blood Transfusion Rates | No significant decrease in the percentage of patients with a postoperative blood transfusion |  |
|  |  |  | Postoperative complications | No advantageous effect was found on postoperative complications |  |
| Calleja, 2016 | IV FCM (Ferinject™)<br>vs.<br>Oral iron | 1) Hgb<12 g/dL in women<br>Hgb <13 g/dL in men<br>2) Serum ferritin <30 ng/mL<br>AND/OR<br>3) Transferrin saturation (TSAT)<br><20% | Hgb levels | Mean total Hgb increases significantly favor the FCM group between diagnosis and hospital admission (1.5 vs. 0.5 g/dL; p<0.0001) and between diagnosis and 30 days post-surgery (3.1 vs. 1.5 g/dL; p<0.0001) | Preoperative treatment in anemic colon cancer patients reduced RBC transfusion requirements and length of hospital stays. |
|  |  |  | Blood Transfusion Rates | Less patients required RBC transfusion during study in FCM group: 9.9% in treatment group vs. 37% in control group (OR: 5.9, 95% CI: 2.9-11.1, p<0.001) |  |
|  |  |  | Iron parameters (Serum ferritin, TSAT) | At 30 days after-surgery, the average FCM (Ferinject™) treated patient presented no recognizable signs of iron deficiency anemia (being the mean of Hgb: 12.6 g/dL [≥12 g/dL]; serum ferritin: 218 ng/mL [≥30 ng/mL]; and saturation transferrin index: 25.1 % [≥20 %]) compared with the no-IV group that did not reach normalized mean values |  |
|  |  |  | Postoperative complications | A lower number of re-interventions and post-surgery complications were seen in the FCM (Ferinject™) group (20.7 vs. 26.5 %; p=0.311) |  |
|  |  |  | Postoperative length of hospital stay | FCM (Ferinject™) group had a significantly shorter mean length of hospital stay (Mean±SD; 8.4±6.8 days) compared to no-IV iron group (Mean±SD; 10.9±12.4 days) |  |
| Kangaspunta, 2022 | IV FCM (Ferinject™)<br>vs. | Hgb<12 g/dL in women<br>Hgb <13 g/dL in men | Blood Transfusion Rates | No significant difference in number of transfusions | Overall decrease in postoperative complications, and postoperative |
| | No IV iron | | Postoperative anemia | Patients treated with IV iron had lower prevalence of anemia at 1 month after surgery (38.7% vs. 65.3%, $p<0.01$) when compared with patients without IV iron treatment. | anemia. |
| | | | Postoperative complications | Patients treated with IV iron had less post-operative complications (33.9% vs. 45.9%, $p=0.045$) | |
|  |  |  | 30-day and 90-day mortality | No significant difference in mortality |  |
|  |  |  | Postoperative length of hospital stay | No significant difference in length of hospital stay |  |
| Titos-Acros, 2012 | Postoperative IV iron III saccharose vs.<br>No postoperative IV iron | Hgb <11 g/dL for all patients | Hgb levels | Mean Hgb at discharge in the IV iron group was $10.0 \pm 1.1$ g/dL vs. $10.6 \pm 1.2$ g/dL in control ($p=0.012$). | IV iron did not appear to impact blood transfusion requirements |
|  |  |  | Blood Transfusion Rates | No significant difference in blood transfusion needs |  |
| Jeong, 2014 | IV IS (Venofer®) vs.<br>No specific treatment | Hgb <10 g/dL for all patients | Hgb levels | Mean increase of Hgb at 6 months post-operation was 3.2 g/dL in the IV iron sucrose group, and 2.5 g/dL in the no-iron group ($P=0.029$). | IV IS (Venofer®) significantly increased serum Hgb in patients developing acute postoperative anemia after gastrectomy |
| Verhaeghe, 2017 | All study patients received IV FCM (Ferinject™) | Serum ferritin < 100 µg/L and TSAT < 20% suggested AID | Hgb levels | Median Hgb change was an increase of 0.5 [IQR; -0.1-1.6] g/dL | Based on the findings, the median increase in Hgb over a 28-day period was 0.5 g/dL. This is not a significant difference but may be due to the inadequate dosing schemes. The response to IV iron depends on the underlying mechanism |
|  |  |  | Iron parameters (Serum ferritin, TSAT) | Median serum ferritin change was an increase of 464 [IQR; 230-830] µg/L<br>Median TSAT change was an increase of 11 [IQR; 5-18.5] % |  |

### Quality Assessment

Three of four randomized trials had a low risk of bias. Keeler et al.[26] had a high risk of bias as patients were not blinded to the study group that they were in. The complete quality assessment of the included RCTs can be found in Figure 2.

**Figure 2.**
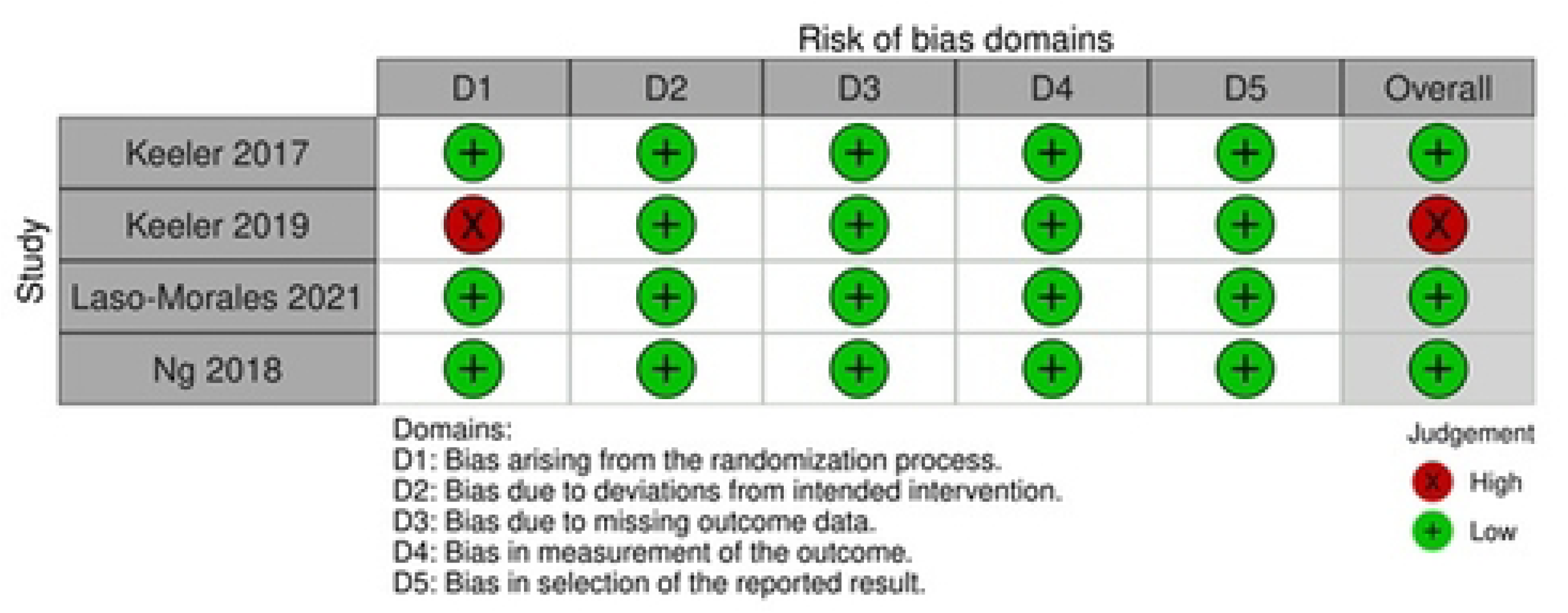
Risk of bias assessment of randomized studies included in the current review.

Of the 13 studies, nine studies had a ‘Moderate’ risk of bias, primarily attributable to appropriately controlled baseline confounding factors, missing data, deviations from intended interventions, and lack of information to assess bias within certain domains. Four studies had a ‘Serious’ risk of bias, likely due to the presence of confounding factors without appropriate statistical considerations, as well as a lack of information. A summary of the risk of bias assessment of non-randomized studies can be found in Figure 3.

**Figure 3.**
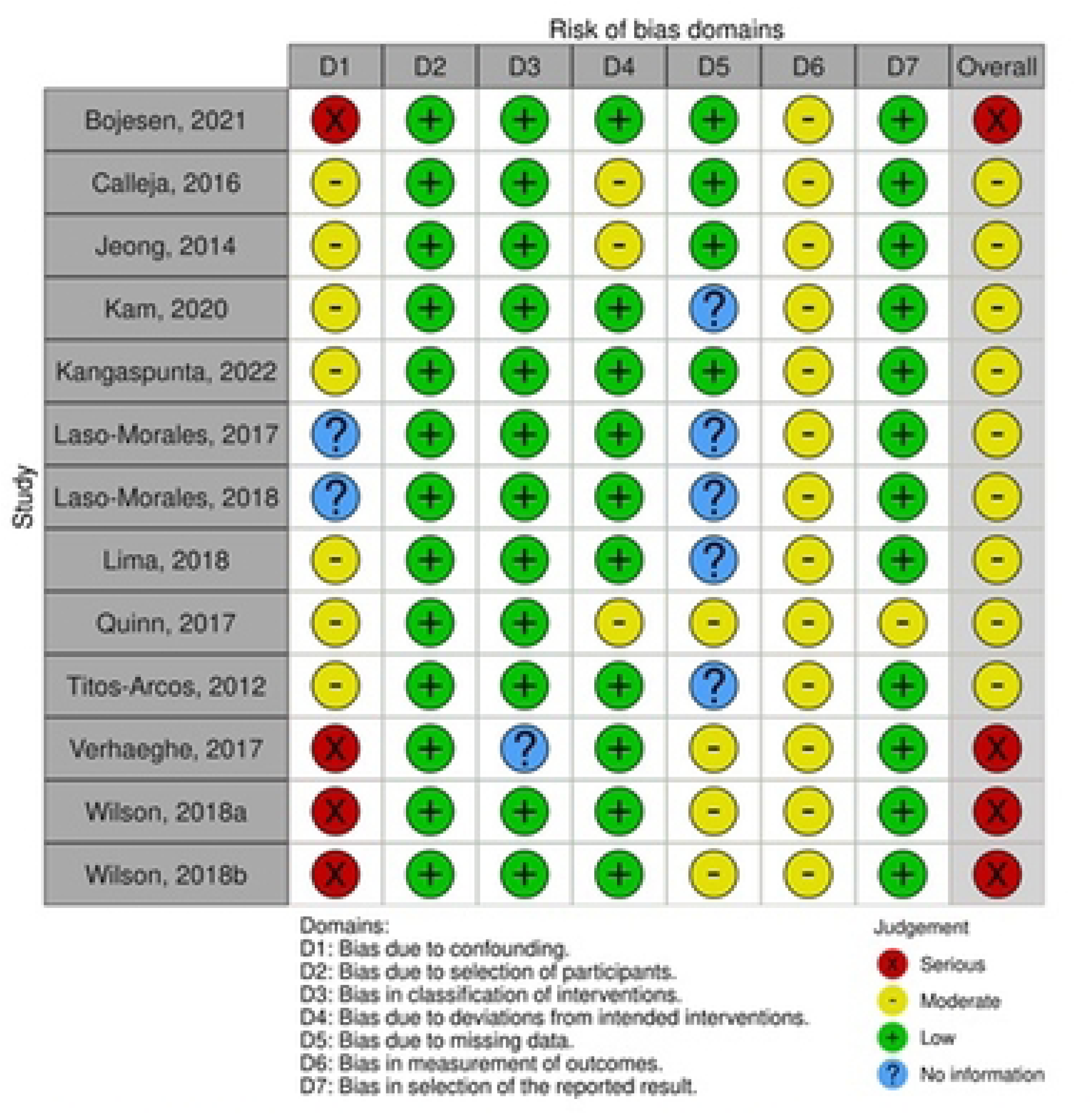
The risk of bias assessment completed for non-randomized studies.

### Outcomes

Key outcomes were determined *a priori* after examination of relevant literature and considerations of the research question. Three key outcomes were identified: Hgb response, RBC transfusion needs, and other applicable health outcomes (e.g., disease free survival (DFS), mortality, morbidity, overall survival (OS), quality of life (QoL), length of hospital stay (LOS), and safety/adverse events).

### Hemoglobin Response

All RCTs except Keeler et al.[26], captured Hgb response. Keeler et al. [25] reported a median increase in Hgb of 1.55 g/dL (IQR; 0.93-2.58) in colorectal cancer (CRC) patients who received 1000 mg or 1500 mg of parenteral FCM (Ferinject™) at least 14 days prior to surgery compared to an increase of 0.5 g/dL (IQR; -0.13-1.33) in patients who received oral ferrous sulfate twice daily until surgery (*p*<0.001). In the investigation conducted by Laso-Morales et al. [27] CRC patients either received a single 1000 mg dose of IV FCM on postoperative day two or 200 mg of IV Iron sucrose (Feriv™) every 48 hours from postoperative day two to discharge. Esophagogastric adenocarcinoma patients in the investigation conducted by Ng et al. [28] received a single 1000 mg dose of iron isomaltoside (Monofer®) prior to the initiation of chemotherapy or received standard of care. Both, Laso-Morales et al.[27] and Ng et al. [28] found no significant differences in Hgb levels between the intervention and comparator groups.

Of the 13 non-randomized studies, ten studies assessed serum Hgb levels. Bojesen et al. [22] reported a mean increase in Hgb of 2.13 g/dL (95% CI: 1.71-2.55 g/dL; *p*<0.0001) after four weeks in CRC patients who received iron isomaltoside prior to surgery. In addition, Lima et al. [23] reported an absolute increase in mean Hgb of 0.9 g/dL (SD 1.3) from baseline to 12-14 weeks (*p*=0.001) in CRC patients who received 1000 mg of IV FCM (Ferinject™) every study visit (12-13 weeks) until anemia or ID was corrected. In contrast, Verhaeghe et al. reported no significant increase in Hgb in patients with GI malignancies who received at least one dose of IV FCM preoperatively after a four week follow up period [24].

Two studies comparing IV iron to oral iron reported an increase in Hgb. Calleja et al. [29] reported a greater increase in Hgb in colon cancer patients who received IV FCM (median dose of 1000 mg given 28.5 days preoperatively) compared to patients who received varying doses and formulations of oral iron (1.5 g/dL vs. 0.5 g/dL; *p*<0.0001) between diagnosis of anemia and hospital admission for surgery, and between diagnosis and 30 days postoperatively (3.1 g/dL vs. 1.5 g/dL; *p*<0.0001). In addition, the percentage of patients with Hgb<10 g/dL was significantly lower in the intervention group at hospital discharge (61.6% vs. 75.7%, *p*<0.05) compared to the oral iron group [29]. Laso-Morales et al. [38] reported a greater increase in Hgb in CRC patients who received IV iron sucrose of varying dose compared to patients who received oral elemental iron (2.0±1.5 g/dL vs. 1.1±1.2 g/dL; *p*=0.001) from postoperative day one to postoperative day 30. However, the prevalence of anemia was greater and more severe in the IV iron group (*p*=0.027) [38].

Jeong et al. [30], Kam et al. [31], Quinn et al. [37], Titos-Acros [33], and Wilson et al. [34] compared IV iron to no specific treatment or standard of care. Jeong and colleagues [30] observed a mean increase of Hgb of 3.2 g/dL in gastric carcinoma patients who received IV iron sucrose every other day in 300 mg doses until total target dosage was given (target dosage calculation can be found in Table 1) compared to an increase of 2.5 g/dL in the no specific treatment group (p = 0.029) six months post-operatively (treatment was initiated post-operatively). Kam et al. [31] reported no difference in mean preoperative Hgb levels in CRC patients who received IV iron or no specific treatment. However, the IV iron groups had a higher median Hgb rise of 1.9 g/dL vs. a rise of 0.6 g/dL in the no specific treatment group (*p*<0.001). In addition, Quinn et al. [37] reported that mean Hgb levels were significantly lower in CRC patients with uncorrected anemia (no treatment or oral iron) compared to patients with corrected anemia (received 1000 mg of IV FCM) on postoperative day three (9.5 g/dL vs. 10.9 g/dL, *p*=0.004). Titos-Acros [33] reported mean Hgb at discharge was lower in colon cancer patients who received 100-200 mg of IV iron saccharose postoperatively, compared to those who did not (10±1.1 g/dL, vs. 10.6±1.2 g/dL; *p*=0.012). Furthermore, Wilson and colleagues [35] reported that patients treated with 1000-2000 mg of IV FCM (Ferinject™) or IV maltoside (Monofer®) less than 6 weeks preoperatively had a significant increase in Hgb compared to the usual care group (1.05 g/dL vs. 0.16 g/dL; *p*<0.001).

### RBC transfusion needs

Three randomized studies reported changes in RBC transfusion needs. Two studies, Keeler et al. [25] and Ng et al. [28], reported no significant differences in blood transfusion needs between IV iron and standard of care. Laso-Morales et al. [32] also reported no significant differences in RBC transfusion needs in colorectal cancer patients. However, it is important to note that this study compared two different IV iron products.

Five non-randomized studies reported changes in RBC transfusion needs. Of the studies that compared IV iron supplementation to oral iron supplementation, Calleja et al. [29] reported that patients in the IV iron group required less RBC transfusions when compared to patients in the oral iron group (9.9% vs. 38.7%; OR: 5.9. 95% CI: 2.9-11.1, *p*<0.001). In addition, Laso-Morales et al. [38] reported higher transfusion needs in the IV iron group compared to the oral iron comparator group (15% vs. 4%, *p*=0.040). Furthermore, Quinn et al. [37] reported that prior to the introduction of the anemia management intervention, anemic patients were 17 times more likely to require perioperative RBC transfusions. In addition, postoperative RBC transfusion rates were 38% in patients who received oral iron or no specific treatment, compared to 0% in patients whose anemia had been corrected by IV iron, and 3.5% in non-anemic patients [37]. Kangaspunta et al. [36] and Titos-Acros [33] reported no difference in RBC transfusion needs between the IV iron group and no IV iron group in their investigations.

### Patient Health Related Outcomes

Three randomized studies collected data on patient health related outcomes. Laso-Morales et al. [27] reported no significant differences between LOS between the IV FCM group and the IV IS group, but did note that the infection rate was lower in the IV FCM group (9.8% vs. 37.2%). Ng et al. [28] reported a marked increase in QoL parameters such as physical and emotional well-being, as well as anemia-specific QoL, with total scores for these indices exceeding the minimum clinically important difference (defined as a difference of one standard deviation from baseline), while no improvement was reported in patients receiving standard of care. In addition, Keeler et al. [26] reported that 11 QoL components (e.g. physical and functional well-being, self-care, pain and disability, general health, etc.) increased by a clinically significant margin in the IV iron group, compared to only one component showing an increase in the oral iron group. Furthermore, patients in the IV iron group had higher median total scores (168, IQR:160-174 vs. 151, IQR:132-170) in the FACT-An than the oral iron group at the time of the outpatient appointment (2-3 months postoperatively) [26].

Six non-randomized studies captured data on various health outcomes. Calleja et al. [29] reported that the IV iron group had a significantly shorter mean length of hospital stay compared to the no-IV iron group (8.4±6.8 days vs. 10.9±12.4 days; *p*<0.001). In addition, Calleja and colleagues [29] reported no adverse events (e.g., deaths, hypersensitivity, or other serious reactions) and there was no difference in post-surgical complications (e.g., suture dehiscence, paralytic ileus, hemoperitoneum, rectal bleeding, thromboembolism, etc.) at 30 days postoperatively [29]. Kangaspunta et al. [36] reported that colon cancer patients treated up to 60 days preoperatively with 500-1000 mg of IV FCM had less post-operative complications (33.9% vs. 45.9%, *p*=0.045), and no difference in LOS, 30- and 90-day mortality between the two groups. Laso-Morales et al. [38] reported no significant differences in postoperative infections, LOS, and complication rates between the two groups, but did report a significantly lower rate of postoperative infection in patients receiving IV iron compared to standard care patients (18% vs. 29%; *p*=0.018). Quinn et al. [37] reported morbidity rates as similar across all groups, but did not provide the data. In addition, LOS was longer for patients with uncorrected anemia compared to patients with corrected anemia (13.2 days vs. 7.2 days; *p*=0.019) [37]. Wilson et al. [34] treated colorectal cancer patients with 1000-2000 mg of IV FCM (Ferinject™) or iron (III) maltoside (Monofer®) and found no significant difference in 1-, 3-, and 5-year OS between the IV iron and non-IV iron groups.

## Discussion

Each included study was used to draw conclusions about the following key findings: Hgb levels, RBC transfusion needs, iron parameters and patient QoL.

A systematic review by Jones *et al.* reveals an improvement in Hgb levels when anemic surgical patients are treated with IV FCM (Ferinject™) [39]. All 10 RCT studies analyzed in the review found an improvement in Hgb concentration from baseline to the end of the study in both the preoperative FCM (Hgb concentration increase from 1.3 g/dL to 4.7 g/dL) and postoperative FCM setting (Hgb concentration increase from 1.7 g/dL to 3.2 g/dL) [39]. Moreover, a retrospective study by Cancado *et al*., evaluated the effects of administering IV iron sucrose (IS; Venofer®) infusions in an IDA (as defined by WHO guidelines) patient population by providing patients with a weekly dose of 200 mg IS until patients received a total iron dose (calculated by weight and Hgb levels of the patient) or when they had a Hgb concentration of greater than 14.0 g/dL [40]. Hgb concentration in patients increased significantly between the baseline and end of study (Mean change: 3.29 g/dL (women) and 4.58 g/dL (men)) [40]. These results coincide with the findings of this review as most of the studies show a significant increase in Hgb levels in patients receiving IV iron treatment opposed to other iron supplementation methods. These findings reveal the strong efficacy of IV iron in effectively increasing Hgb levels in patients with IDA.

Hallet *et al*., systematically reviewed four studies to assess the effects of perioperative iron supplementation on RBC transfusion needs in patients undergoing elective GI surgeries [41]. The study found that although fewer patients required transfusions when given iron supplementation, the observations were statistically insignificant [41]. However, the findings of their systematic review may be biased, as the results were based on four studies with small sample sizes which may not provide an accurate effect estimate of iron supplementation [41]. Based on our systematic review findings, most studies that evaluated RBC transfusion rates as an outcome of interest found no significant difference between groups. However, a multicenter cohort study by Calleja *et al*. found that patients in the group receiving IV FCM (n= 111) needed less RBC transfusions than patients receiving oral iron supplementation (n= 155) (OR: 5.9, 95% CI: 2.9-11.1, p<0.001) [29]. Therefore, additional studies need to be conducted to assess the impact of IV iron and comparator iron types on blood transfusion rates to validate and strengthen the current evidence available. Furthermore, valid clinical endpoints, such as Hgb rise, effective iron repletion, less RBC transfusion, morbidity, QoL and hospital length of stay for inpatient assessments, should be assessed in these studies.

Iron parameters were another key factor explored in studies evaluating the efficacy of IV iron treatment in IDA correction. Jones *et al*. found that in the preoperative setting, IV iron intervention revealed a 15-35% increase in TSAT levels from baseline and an increase in serum ferritin levels from 19 µg/L at baseline to 229-558 µg/L [39]. Additionally, they found that in the postoperative setting, there was a 7.2-20% increase in TSAT and a serum ferritin increase from 19 µg/L to 114-571 µg/L [39]. In our review, seven studies assessed iron parameters (serum ferritin and TSAT) to evaluate the difference in these biomarkers after treating patients with IV iron. All studies found a statistically significant increase in ferritin and TSAT levels of patients receiving IV iron compared to comparator groups. However, a limitation to these study findings is the use of serum ferritin as an indicator of IDA improvement in cancer patients. Ferritin levels are found to be elevated in the cancer patient population due to the cancer’s inflammatory nature [42]. The elevated levels of serum ferritin in cancer patients could be due to the abnormal production and release of ferritin from tumour cells [43]. Thus, the sensitivity of ferritin as a prognostic value for iron deficiency is low and studies should refrain from using this parameter as an indicator of iron levels in cancer patients.

QoL is an important factor that is often overlooked when treating IDA and assessing patient performance in an oncology population [29]. A review written by *Strauss and Auerbach* evaluated the importance of using validated patient reported outcome tools (FACT measurement system) to assess QoL (34). They found IV iron (IV ferric gluconate) to be the most effective treatment for IDA in cancer patients which resulted in an improvement in patient FACT scores (34). One RCT assessed in their review by Henry *et al*. looked at the impact of IV iron on FACT-Fatigue scores in anemic patients receiving chemotherapy [29]. All patients in the trial received epoetin alfa once a week for four weeks, then an adjusted dose based on their protocol [29]. These patients were randomized into three groups: no iron (ESA alone), oral iron sulfate with ESA, or IV ferric gluconate with ESA [29]. Researchers found that patients receiving IV ferric gluconate reported a significant improvement in the FACT-Fatigue scale (MID=3) compared to patients receiving oral iron or no iron [29]. In our systematic review, we found that the information on QoL specifically in GI cancer patients was limited. Only two studies evaluated QoL as an outcome of interest. Keeler *et al*., found that the QoL Fact-An scores were higher in patients receiving IV iron compared to oral iron (FACT-An total score (oral iron 151 (132–170 [69–183]); IV iron 168 (160–174 [125–186]); p = 0.005))) [26]. However, these conclusions may not be meaningful due to the small sample size (n = 116) of the study [26]. More QoL research should be conducted in the GI cancer population to gain a stronger understanding of QoL improvement when given iron supplementation.

The strengths of this systematic review include the comprehensive and inclusive search strategy that was approved by an oncologist specializing in GI cancers, as well as a hematologist. This review assessed a total of 1,623 studies across five databases with no restrictions to language or publication types. Two authors (SN and NS) independently screened the eligibility of each of the studies to minimize selection bias. Moreover, the two authors (SN and NS) conducted separate data extractions for studies that passed the initial screening phase to ensure all pertinent information was collected and all studies followed the eligibility criteria of the review. Furthermore, the quality of both RCTs and non-RCTs were assessed using Cochrane’s RoB 2 and ROBINS-I tools. The risk assessment revealed the limitations of the studies included in this systematic review and helped curate a thorough understanding of the missing findings in current publications.

This systematic review presents some limitations. Due to the varying interventions, comparators, populations, clinical endpoints, iron formulation, dosing schemes and settings evaluated in each RCT, the results of the review produced heterogeneous findings and thus, made it difficult to conduct a meta-analysis. Additionally, the studies assessed in this review used different time points of data collection, included patients undergoing different surgical approaches, and different cancer types, making it difficult to make valid comparisons between findings. Some studies also assessed outcomes that did not provide meaningful conclusions, such as the use of serum ferritin as a prognostic parameter in this patient population. Moreover, a majority of studies in this review were potentially underpowered due to their small sample sizes.

Overall, to strengthen the findings in this field, research should be conducted using larger sample sizes to validate findings prior to making conclusive statements on the efficacy of the intervention being used. Additionally, research evaluating the efficacy of IV iron should use iron parameters that are solely influenced by iron intake in IDA patients (eg. Hgb levels, RBC transfusion rates, serum iron, and TSAT) to ensure findings are resultant of the type of iron patients receive opposed to co-factors such as chemotherapy. More studies should also follow patients for a longer duration of time to assess the long-term impacts of IV iron on QoL and other important outcomes such as survival in this patient population. Another important consideration future researchers should incorporate in their research is the drug availability of IV iron compared to oral iron and how physicians can implement an IV iron infusion center in a feasible manner, especially as there are newer IV iron formulations (e.g. ferric derisomaltose) requiring shorter infusion times.

The findings of this systematic review reveal the importance of addressing the prevalence of IDA in a GI oncology patient population. Patients should receive timely diagnosis and management of ID prior to undergoing cancer treatments to avoid postoperative complications related to anemia, including myelosuppressive chemotherapy dose reductions, and to improve overall QoL. More research in this field will help create an international set of guidelines to ensure best clinical practices to improve IDA in patients with GI malignancies.

## Data Availability

All relevant data are within the manuscript and its Supporting Information files.

## Appendix

### Full Search Strategy (no restrictions)

> (functional iron deficiency OR functional iron deficient anemia OR absolute iron deficiency OR absolute iron deficient anemia OR cancer related anemia OR chemotherapy-induced anemia OR chemotherapy induced anemia OR radiotherapy induced anemia OR radiotherapy-induced anemia OR anemia OR anaemia OR iron deficien* anemia OR cancer-related anemia OR microcytic anemia) AND (treatment OR therapy OR chemotherapy) AND (gastric cancer OR gastroesophageal cancer OR gastrointestinal cancer OR adenocarcinoma OR gastr* neoplasm OR gastr* carcinoma OR colo* cancer OR colo* neoplasm OR colo* carcinoma OR stomach cancer OR stomach neoplasm OR stomach carcinoma) AND (ESA OR ESA therap* OR erythropoiet* stimulating agents OR intravenous iron OR IV iron OR iron replacement OR iron supplement* OR parenteral iron OR iron sequestration OR myelosuppression OR iron studies OR serum iron OR total iron binding capacity OR transferrin saturation)

## Acknowledgments

This research did not receive any specific grant from funding agencies in the public, commercial, or not-for-profit sectors.

## Declarations

### Author Contributions

*Shankavi Nandakumar*: Literature search, figures, study design, data collection, data analysis, data interpretation, writing, conceptualization, data curation, investigation, methodology, project administration, resources, validation, visualization, writing - original draft, writing - review and editing

*Navreet Singh*: Literature search, figures, study design, data collection, data analysis, data interpretation, writing, conceptualization, data curation, investigation, methodology, project administration, resources, validation, visualization, writing - original draft, writing - review and editing

*Alliya Remtulla Tharani*: Conceptualization, project administration, supervision, writing - review and editing

*Christine Brezden-Masley*: Conceptualization, project administration, supervision, writing - review and editing

Note: Shankavi Nandakumar and Navreet Singh contributed equally.

### Data Availability Statement

Data sharing is not applicable to this review article.

